# Unravelling the determinants of human health in French Polynesia: the MATAEA project

**DOI:** 10.1101/2023.04.05.23288204

**Authors:** Iotefa Teiti, Maite Aubry, Sandrine Fernandes-Pellerin, Etienne Patin, Yoann Madec, Pauline Boucheron, Jessica Vanhomwegen, Jérémie Torterat, Stéphane Lastère, Sophie Olivier, Anthony Jaquaniello, Maguelonne Roux, Vincent Mendiboure, Christine Harmant, Aurélie Bisiaux, Gaston Rijo de León, Dang Liu, Hervé Bossin, Françoise Mathieu-Daudé, Clémence Gatti, Edouard Suhas, Kiyojiken Chung, Bertrand Condat, Pierre Ayotte, Nicolas Prud’homme, Eric Conte, Nathalie Jolly, Jean-Claude Manugerra, Anavaj Sakuntabhai, Arnaud Fontanet, Lluis Quintana-Murci, Van-Mai Cao-Lormeau

## Abstract

**Background:** French Polynesia is a French overseas collectivity in the Southeast Pacific, comprising 75 inhabited islands across five archipelagoes. The human settlement of the region corresponds to the last massive migration of humans to empty territories, but its timeline is still debated. Despite their recent population history and geographical isolation, inhabitants of French Polynesia experience health issues similar to those of continental countries. Modern lifestyles and increased longevity have led to a rise in non-communicable diseases (NCDs) such as obesity, diabetes, hypertension, and cardiovascular diseases. Likewise, international trade and people mobility have caused the emergence of communicable diseases (CDs) including mosquito-borne and respiratory diseases. Additionally, chronic pathologies including acute rheumatic fever, liver diseases, and ciguatera, are highly prevalent in French Polynesia. However, data on such diseases are scarce and not representative of the geographic fragmentation of the population.

**Objectives:** The MATAEA project aims to estimate the prevalence of several NCDs and CDs in the population of the five archipelagoes, and identify associated risk factors. Moreover, genetic analyses will contribute to determinate the sequence and timings of the peopling history of French Polynesia, and identify causal links between past genetic adaptation to island environments, and present-day susceptibility to certain diseases.

**Methods:** This cross-sectional survey is based on the random selection of 2,100 adults aged 18-69 years and residing on 18 islands from the five archipelagoes. Each participant answered a questionnaire on a wide range of topics (including demographic characteristics, lifestyle habits and medical history), underwent physical measurements (height, weight, waist circumference, arterial pressure, and skin pigmentation), and provided biological samples (blood, saliva, and stool) for biological, genetic and microbiological analyses.

**Conclusion:** For the first time in French Polynesia, the MATAEA project allows to collect a wide range of data to explore the existence of indicators and/or risk factors for multiple pathologies of public health concern. The results will help health authorities to adapt actions and preventive measures aimed at reducing the incidence of NCDs and CDs. Moreover, the new genomic data generated in this study, combined with anthropological data, will increase our understanding of the peopling history of French Polynesia.

## INTRODUCTION

French Polynesia is located in the Southeast Pacific and within the Polynesian triangle, which is bounded by the islands of Hawaii to the North, Aotearoa (New Zealand) to the West and Rapa Nui (Easter Island) to the East. This French overseas collectivity extends over a maritime area as large as Europe and includes 121 islands, of which 75 are inhabited, distributed within five archipelagoes: Society, Marquesas, Tuamotu, Gambier and Austral (Figure 1). According to the previous national population census conducted in 2017, there are about 276,000 inhabitants distributed in the various archipelagoes, of which nearly 75% are concentrated in the islands of Tahiti and Moorea (Windward Islands, Society archipelago) (1).

**Figure 1.**
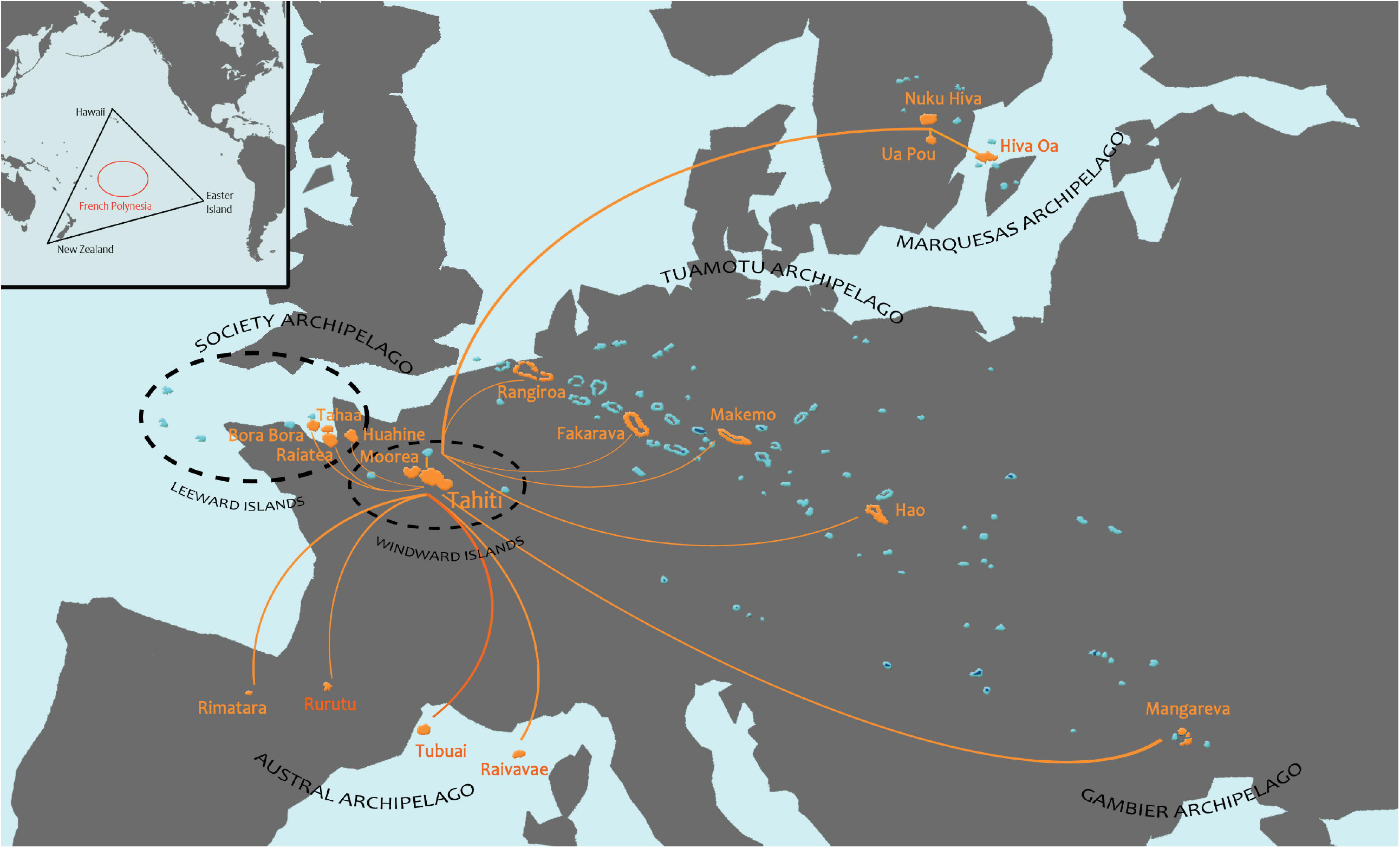
Map of French Polynesia showing the geographical distribution of the five archipelagoes (Society, Tuamotu, Gambier, Marquesas and Austral) compared to the European continent (showed at the same scale). Flight connections between Tahiti and the islands selected for the survey are indicated by orange lines. Inset map at upper left shows location of French Polynesia (red circle) within the Polynesian triangle (black triangle) bounded by the islands of Hawaii, New Zealand and Easter Island in the Pacific Ocean.

In the course of the history of human migrations around the world, Pacific islands, and Polynesia in particular, were the last empty lands to be settled (2). Although archaeological and linguistic data have provided evidence for the geographical and temporal origin of the populations of the Eastern Pacific islands, the sequence and timings of settlement of the Polynesian islands are still debated (3). To help elucidate these gaps, a few genetic studies including samples from Pacific populations have been conducted over the past five years (2-6). However, for the studies involving the use of samples from the population of French Polynesia, only a limited number of samples collected in a few islands were available. Some studies also aimed to identify genomic variations related to adaptation of the populations to island environments. In a study including genomes from 20 populations from the Pacific region, candidate variants for positive selection were found in genes relating to immunity and metabolism, suggesting genetic adaptation to pathogens and food sources specific to Pacific islands (4). However, further investigations are needed to identify causal links between past genetic adaptation and present-day disease risk in Pacific settings. Indeed, most studies investigating genetic links with vulnerabilities to non-communicable (NCDs) and communicable (CDs) diseases have focused on populations in Africa, Europe, Asia, and USA (7-10). The same observation is made for the research investigating the link between human gut microbiome composition and susceptibility to diseases (7). Despite the high burden of various NCDs and CDs, Pacific islands remain highly underrepresented in genomic studies.

In addition to genetic and microbial factors, social and environmental factors such as poverty, education, and access to healthcare are thought to be major drivers of disease burden in the Pacific islands (7). In French Polynesia, prevention measures against NCDs and CDs have been enhanced, and access to healthcare has been improved in all archipelagoes over the past 3 decades (11). This has certainly contributed to the extension of life expectancy at birth, which has increased from 67.1 years to 74.7 for men and 72.8 years to 79.1 for women between 1990 and 2020 (12). However, modern lifestyles (including overconsumption of food and physical inactivity) and increased longevity have led to a rise in the incidence of NCDs (13). The most recent data collected in French Polynesia in 2010 indicated that the prevalence of obesity and hypertension in adults was 40.4% and 26.7%, respectively. It was estimated that only 1.7% of the population of French Polynesia presented a low risk of developing a NCD whereas 45% were at high risk (14). According to the latest reports from the health authorities of French Polynesia, the leading causes of death are cardiovascular diseases (26.1%) mainly associated with lifestyle, and cancers (25.1%) (15, 16). Among the Pacific island countries and territories (PICTs), French Polynesia displayed one of the highest age-standardized cancer incidence (267.9 for males and 214.0 for females per 100,000) and age-standardized cancer mortality (157.2 for males and 106.8 for females per 100,000) in 2020 (17-19). The most frequently reported cancers are lung, prostate, breast, thyroid and colorectal cancers (19). Moreover, the incidence of hepatocellular carcinoma recorded in the Austral archipelago between 2013 and 2017 is among the highest in the world (43.1/100,000 inhabitants), this pathology being associated with a high prevalence of infection with hepatitis B virus (HBV) and obesity (20).

Dietary habits, especially high fish consumption, also expose the population of French Polynesia to seafood-related NCDs. Among them, ciguatera poisoning, a disease caused by the consumption of reef fishes or marine invertebrates contaminated by neurotoxins (ciguatoxins) produced by the microalgae *Gambierdiscus*, is highly prevalent in French Polynesia where it constitutes an important public health issue (21). In 2021, the overall incidence rate of ciguatera, based on the number of symptomatic cases reported by healthcare workers or by the patients themselves, was 6.9/10,000 inhabitants (22). However, as (i) ciguatera is not on the list of notifiable diseases, (ii) a large proportion of ciguatera-affected people do not consult a healthcare worker, and (iii) cases requiring emergency care or hospitalization in the main hospital of French Polynesia (*Centre hospitalier de Polynésie française*, Tahiti) are not systematically recorded, the true incidence rate of this disease is underestimated. Another source of seafood-related contaminants are heavy metals including mercury and lead. Indeed, high levels of mercury were detected in adolescents and adults from two atolls of the Tuamotu archipelago (Hao and Makemo) (23), in adults from the Leeward islands (Tahiti and Moorea) (24), and in cord blood samples of delivering women from all archipelagoes, suggesting that Polynesian newborns may be prenatally exposed to high doses of mercury (25). Moreover, concentrations of lead exceeding toxicological reference values were found in a significant proportion of the residents of Hao and Makemo islands tested to assess their exposure to toxic metals and polychlorinated biphenyls (PCBs) (23). Although levels of PCBs found in both atolls were low compared to international standards, there was a higher body burden of PCBs in residents of Hao which was suggested to be potentially linked to some contamination of the environment by military support activities in this island. Another source of environmental contamination that may have an impact on the population health are pesticides. To our knowledge, there are no data concerning the level of exposure to pesticides of the population of French Polynesia. However, surveys on the pollution of French Polynesia coral reefs by pesticides found the presence of herbicides and insecticides in fishes which are commonly consumed by the local inhabitants (26-28).

In addition to NCDs, CDs represent major health problems in French Polynesia. As a result of the prevention, screening, treatment and vaccination programs implemented by the health authorities, the incidence of several CDs has decreased (29). Despite these efforts, some CDs caused by bacterial infections persist, including leprosy (mean incidence of 1.6 cases per 100,000 inhabitants reported from 2000 to 2017) and tuberculosis (incidence of 19.4 cases per 100,000 inhabitants in 2018) (30, 31). Moreover, in 2013, the prevalence of Bancroftian lymphatic filariasis, an infection caused by the nematode worm *Wuchereria bancrofti* transmitted by mosquitoes, still had not reached the elimination threshold (*i*.*e*. filarial antigenemia <1%) defined by the World Health Organization (WHO) in Tahiti, Huahine (Leeward Islands, Society archipelago) and three islands in the Marquesas archipelago (Tahuata, Hiva Oa and Fatu Hiva), despite several years of community treatment (32-35). Furthermore, HBV remains a public health problem, despite the availability of an effective vaccine. Following the finding of hepatitis B surface antigens (HBsAg) among 10.48% inhabitants of the Austral archipelago randomly selected in 1987 (36), vaccination of children at birth started to be implemented in 1992 to decrease the burden of chronic HBV infection in French Polynesia (37), and became mandatory in 1995 (38). A nationwide serosurvey conducted in 2014 in 6 years old schoolchildren from the five archipelagoes showed a three-dose vaccination coverage of 98% and a prevalence of HBsAg of 0% (39). However, screening in 2019 of all adult residents of Rapa, the most isolated island of the Austral archipelago, revealed a prevalence of HBsAg of 11% in people born before 1992 (20).

Among the CDs considered as important public health issues in French Polynesia, those caused by arthropod-borne viruses (arboviruses) represent the highest risk for the population. Indeed, the four serotypes of dengue virus (DENV-1 to -4) have caused multiple outbreaks over the last decades, with deaths frequently reported in children (40, 41). Moreover, Zika virus (ZIKV) and chikungunya virus (CHIKV) caused explosive outbreaks in 2013-2014 (42) and 2014-2015 (43), respectively. During the ZIKV outbreak, severe neurologic complications in adults (44) and central nervous system malformations in newborns and fetuses (45) were recorded. In addition to arboviral diseases, CDs caused by respiratory viruses have become major health concerns over the past decades. Several epidemics of variable severity caused by influenza A and/or B viruses have been reported in French Polynesia (29). While influenza was not considered a public health priority until the early 2000s, an epidemic caused by the influenza A virus subtype H1N1 highly affected the population of French Polynesia in 2009, with 13 severe cases requiring hospitalization in a critical care service and 7 deaths recorded (46). More recently, the severe acute respiratory syndrome coronavirus 2 (SARS-CoV-2) reached French Polynesia in March 2020 and caused several epidemic waves, with a total of 78,864 confirmed cases, 3,315 hospitalizations and 783 deaths reported as of August 31, 2022. A serosurvey conducted at the end of 2021, from a representative sample of the adult population of Tahiti, found that 57.7% of the study participants had been infected by SARS-CoV-2 during the first two epidemic waves (47).

Bacterial infections also represent a significant risk for the population of French Polynesia. The incidence of leptospirosis, a rodent-borne disease caused by bacteria belonging to the order *Spirochaetales*, exploded in 2017 with a rate of 72/100,000 inhabitants (48). Moreover, the incidence of *Staphylococcus aureus* (*S. aureus*) infections was estimated at 38/100,000 inhabitants in 2008, and this bacterium was shown to be responsible for one third of the community bacterial infections documented in French Polynesia. The proportion of *S. aureus* strains resistant to methicillin increased from less than 10% in 1995 to more than 35% in 2001 (49). French Polynesia is also characterized by the unusual high prevalence of acute rheumatic fever associated with infection by Group A streptococcus, with an incidence of 0.06% and a prevalence of 1.29% reported in 2015 (29, 50).

Although data exist on the levels of exposure of the population and the risk factors associated with NCDs and CDs in French Polynesia, these data are often old, fragmentary and concern a few islands. Moreover, genomic data, based on whole-genomes, from the Polynesian populations are lacking, precluding a comprehensive assessment of the settlement history of the various islands and archipelagoes, and of the history of adaptation of these populations to island environments - an information that should be of great benefit for the study of the genetic basis of certain diseases that present high incidence in the region. Here we describe the MATAEA project, a wide cross-sectional population-based survey designed to investigate the determinants of health in the adult population of French Polynesia, using a combination of data obtained from a questionnaire, physical measurements, and biological analyses in 2100 inhabitants of 18 different islands distributed among the five archipelagoes.

## STUDY OBJECTIVES

MATAEA is a Tahitian compound word (“MATA” means “eye” and “EA” means “health”) which can be translated by “take a look at health”. The first objective of the MATAEA project was to collect a wide range of data from a large number of inhabitants to assess the health status of the adult population of the five French Polynesian archipelagoes, and identify risk factors for NCDs and CDs related to the specific living context (including the lifestyle, place of residence, history of infections) and intrinsic characteristics of individuals (such as genetics, age, sex, oral and gut microbiome). Results will be made available to health authorities for them to adapt actions and preventive measures to better reduce diseases incidence. The second objective of the project was to use genomic data obtained from the residents to increase our understanding of the history of the settlement of the French Polynesian islands, and to identify the major demographic and adaptive events that may explain the current incidence of some NCDs, and the higher susceptibility to certain NCDs and CDs. Although designed as a general-purpose survey with a broad scope, topics identified as major public health issues will be given priority treatment by specialized laboratories involved in the project, as described below.

First, prevalence data for metabolic diseases (including obesity and diabetes) dating from 2010 will be updated (14), and their associated risk factors (such as dietary habits, physical activity, blood pressure, high blood sugar, excess body fat around the waist, abnormal cholesterol or triglyceride level, genetic susceptibility factors, microbiome composition) will be investigated.

Second, the seroprevalence of mosquito-borne diseases known to have previously circulated in French Polynesia (including the four serotypes of dengue, Zika and chikungunya) will be measured as previously published (51, 52), to assess the risk for new outbreaks to occur. In addition, the prevalence of filarial antigenemia will be evaluated to check whether the elimination threshold recommended by the WHO has been reached in all archipelagoes, subsequent to the strengthening of community treatment in islands most affected by Bancroftian lymphatic filariasis (35). Risk factors for those mosquito-borne diseases will also be sought. Among them, the level of exposure of the population to mosquito bites will be measured by analyzing the presence of antibodies targeting mosquito salivary proteins in the blood of participants, as previously described (53).

Third, screening for HBsAg and anti-HBV antibodies will be performed to assess the prevalence of current and past HBV infections among the inhabitants of the five archipelagoes, especially those with increased risk factors such as being borne before 1995 and living in the Austral archipelago. As there is no available data regarding the prevalence of hepatitis C virus (HCV) infections in French Polynesia, the presence of anti-HCV antibodies will also be tested.

Fourth, concentrations of several heavy metals (including mercury and lead) and pesticides will be measured in the blood of inhabitants of the five archipelagoes to assess the degree of environmental contamination and the risk for human health. Data collected in the questionnaire, such as the island of residence and dietary habits, will be used to identify risk factors associated with contamination with toxic metals and pesticides, as well as ciguatera poisoning. As there is no confirmatory test available to detect current or past ciguatera poisoning, estimate of the prevalence of this disease in the population of the five archipelagoes will be based on the information self-reported by the participants in the questionnaire.

Fifth, as SARS-CoV-2 was introduced into French Polynesia during the participants’ inclusion period and a large number of cases with mild or no symptoms might have not been reported, the presence of specific antibodies will be analyzed in the blood samples of participants to evaluate the real proportion of the population infected in the different archipelagoes after the emergence of the virus.

Sixth, serological analyzes will be conducted for a panel of viruses known to regularly cause infections in French Polynesia (such as influenza viruses, human immunodeficiency virus, rotaviruses and enteroviruses), but for which there is little data on the immune status of the population.

Finally, the seroprevalence of rodent-borne pathogens including Hantaviruses will be determined, as the high incidence of leptospirosis suggests that other diseases transmitted by rodents may circulate in French Polynesia. The only serosurvey of hantavirus infections in French Polynesia was conducted in 1989 and found specific antibodies in a few samples from rats and humans (54). Consequently, data collected in the present study will allow to detect possible silent circulation of Hantaviruses.

## MATERIALS AND METHODS

### Sampling Design

The MATAEA project is a cross-sectional population-based survey conducted on a random sample of the general population aged from 18 to 69 years, representative of the five archipelagoes of French Polynesia. The sampling protocol is based on the previous population census dating from 2017 (1). A sample with a target size of 2100 participants, equally divided into three age groups (18-29, 30-44 and 45-69), with a sex ratio of 1:1, was established. To represent all archipelagoes, French Polynesia was stratified into three geographical strata: Windward Islands (Society archipelago), Leeward Islands (Society archipelago), and other archipelagoes (Tuamotu, Gambier, Marquesas and Austral), which respectively include 75%, 13% and 12% of the population (Table 1). Since the population size in the Windward Islands is much higher than in the Leeward Islands and in the other archipelagoes, the sample size was amplified in the latter two strata to keep sufficient precision around prevalence estimates for the health conditions under consideration. Consequently, the same sample size of 700 subjects was chosen for the three strata, allowing a precision never exceeding +/- 3.7% around prevalence estimates within each strata. In each strata, for convenient reasons, the most populated islands with a health care center and an airport with regular flight rotations were selected for the study, and the number of participants to be recruited was proportional to the population size of the islands as detailed in Table 1. In each island, households were randomly selected by the *Institut de la Statistique de la Polynésie française* (ISPF, Tahiti), and in each household one resident meeting the criteria of age and sex sought for the study was recruited.

**Table 1.** Geographical distribution of the 2,100 participants randomly selected for the MATAEA project.

| Strata |  |  | Archipelago | Island |  |  |
| --- | --- | --- | --- | --- | --- | --- |
| Name | Population size (%)* | Sample size | Name | Name | Population size (%)* | Sample size |
| Windward Islands | 207,333 (75%) | 700 | Society | Tahiti | 189,517 (91%) | 637 |
|  |  |  |  | Moorea | 17,816 (9%) | 63 |
| Leeward Islands | 35,393 (13%) | 700 | Society | Raiatea | 12,249 (36%) | 252 |
|  |  |  |  | Bora Bora | 10,549 (31%) | 217 |
|  |  |  |  | Tahaa | 5,234 (15%) | 105 |
|  |  |  |  | Huahine | 6,075 (18%) | 126 |
| Others archipelagoes | 33,192 (12%) | 700 | Tuamotu | Rangiroa | 3,657 (16%) | 112 |
|  |  |  |  | Fakarava | 1,637 (7%) | 49 |
|  |  |  |  | Hao | 1,258 (5%) | 35 |
|  |  |  |  | Makemo | 1,508 (6%) | 42 |
|  |  |  | Gambier | Mangareva | 1,535 (7%) | 49 |
|  |  |  | Marquesas | Nuku Hiva | 2,951 (13%) | 91 |
|  |  |  |  | Hiva Oa | 2,243 (10%) | 70 |
|  |  |  |  | Ua Pou | 2,213 (9%) | 63 |
|  |  |  | Austral | Tubuai | 2,217 (9%) | 63 |
|  |  |  |  | Rurutu | 2,466 (10%) | 70 |
|  |  |  |  | Raivavae | 903 (4%) | 28 |
|  |  |  |  | Rimatara | 872 (4%) | 28 |
| **Total** | 275,918 | 2,100 |  |  |  | 2,100 |

\* data from the population census of 2017 (59)

### Study timeline

The recruitment of participants in the 18 islands selected for the study was initially planned from November 2019 to June 2020. However, following the introduction of SARS-CoV-2 in French Polynesia in March 2020, several interruptions occurred during the recruitment phases and the sampling period lengthened (Figure 2). The first break was caused by the population lockdown implemented from March 20 to May 21, 2020. Inclusions resumed in June 2020 but stopped the following month due to the occurrence of a first epidemic wave caused by the original Wuhan strain. Inclusions resumed once again in April 2021 but stopped during August and September 2021, because of a second epidemic wave caused by the Delta variant. The last recruitments were performed from October to December 2021.

**Figure 2.**
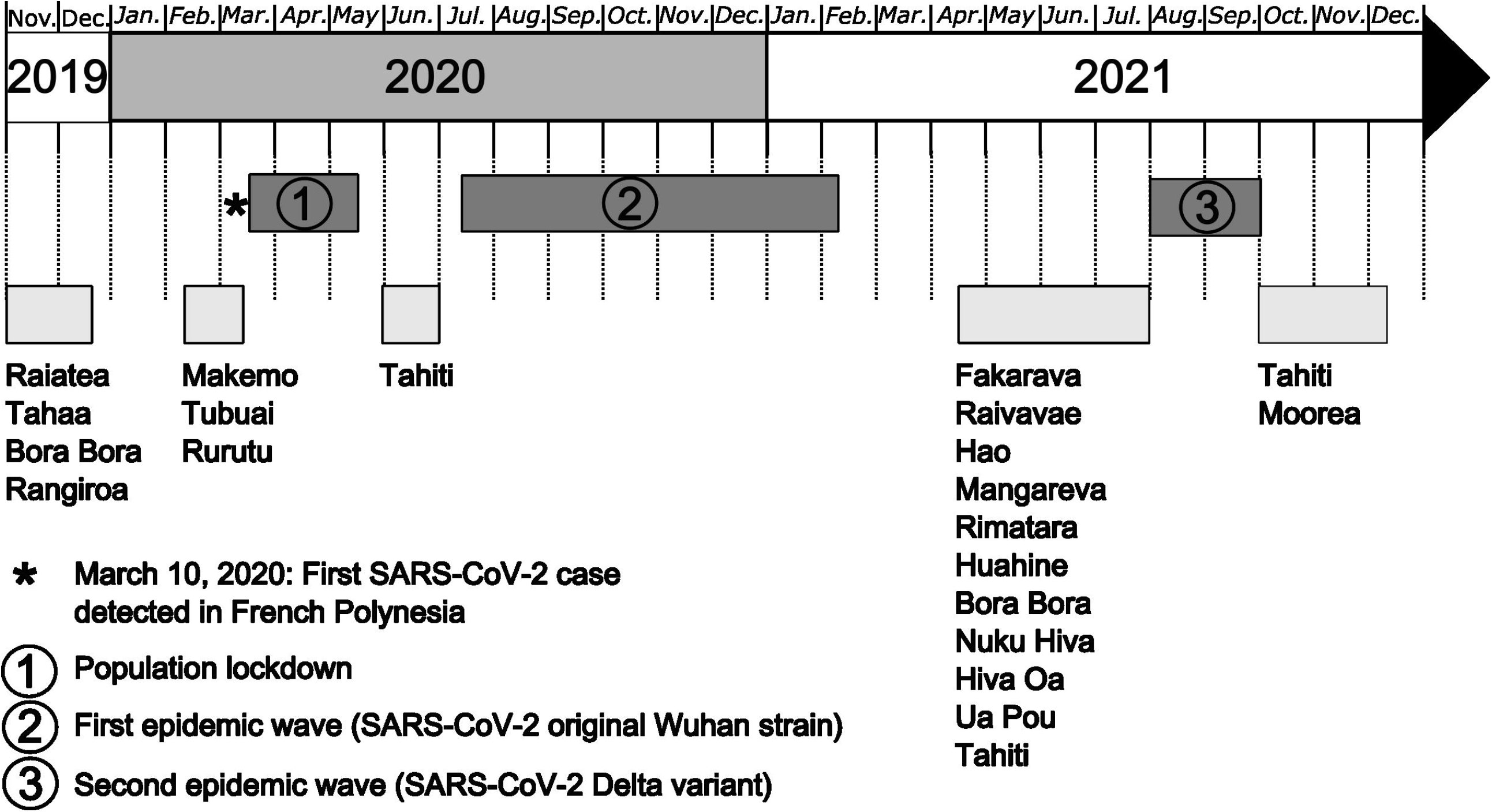
Study timeline for the MATAEA project. Recruitment of participants in the 18 islands of French Polynesia started on November 4, 2019, and ended on December 8, 2021. Three breaks occurred during that period because of SARS-CoV-2 circulation: (1) population lockdown from March 20 to May 21, 2020; (2) first epidemic wave due to the SARS-CoV-2 Wuhan strain from August 2020 to February 2021; (3) second epidemic wave due the SARS-CoV-2 Delta variant from August to September 2021.

### Inclusion/exclusion criteria

The study enrolled men and women aged from 18 to 69 years at the time of enrollment, who had been residing in French Polynesia for at least six months, were affiliated to the French Polynesia social security or assimilated regimens, and were able to provide a written informed consent. Were excluded from the study the minors, pregnant or parturient women, persons deprived of their liberty by a judicial or administrative decision, adults being subject to a legal protection measure or unable to express their consent, people undergoing psychiatric care, people not affiliated to the French Polynesia social security or assimilated regimens, disabled people, and the homeless.

### Participant recruitment and informed consent

Each household randomly selected for the study was visited by trained staff from the *Institut Louis Malardé* (ILM, Tahiti), and only one resident was recruited in the household. Detailed information on the purpose and procedures of the study were provided, to the individual selected, by the ILM staff. Then, signed written informed consent was obtained from the participant. Participation in the study involved agreeing to answer a questionnaire, to undergo physical measurements (height, weight, waist circumference, arterial pressure, and skin pigmentation), and to provide biological samples (blood, saliva, and optionally stool), as detailed in the next sections. The participant answered the questionnaire during face-to-face interview, and answers were reported on an electronic tablet device. Then, the participant was invited to provide a stool sample and was given a collection tube in case of acceptance. Finally, an appointment with a nurse was scheduled one to three days later, in the morning, to collect physical measurements and biological samples. The participant was asked to fast for at least 8 hours and to collect and store the stool sample in a fridge within 24 hours before the visit of the nurse.

Participants received no financial compensation. However, they freely benefited from the results of the anthropometric, blood pressure and fasting blood glucose measurements collected by the nurse, and of the hematology and biochemistry analyses performed from their blood samples at the clinical laboratory of ILM. Moreover, and importantly, participants were informed if any atypical result was detected and were subsequently guided through a visit to a physician.

### Questionnaire

Case report forms (CRF) were built with ODK Collect (Get ODK Inc., USA) which is an open-source mobile client. The CRF consisted of 216 items divided into three major sections: socio-demographic status, lifestyle behaviors, and general health and medical history. It contained questions from the standardized STEPS questionnaire of the WHO designed to monitor the main NCDs risk factors (55), and additional questions specific to the context of French Polynesia. Questions related to COVID-19 were added in the questionnaire in April 2021. The detailed CRF is available in Supplemental Data 1.

### Physical measurements

#### Anthropometric measurements

Anthropometric measurements included height, weight and waist circumference. Standing height was measured in centimeters (cm) with a portable stadiometer (SECA, model 213, France). Weight was measured in kilograms (kg) using a portable electronic weighting scale (SECA, model 813, France). Waist circumference was measured in cm using a constant tension tape (SECA, model 203 measuring tape, France) at the midpoint between the lowest rib and above the iliac crest. Body mass index (BMI) was calculated as weight (in kg) divided by height (in m) squared. BMI values were categorized according to standardized criteria (56).

#### Blood pressure measurements

Blood pressure was measured on the left arm at heart level using an automatic digital blood pressure monitor (Omron model HBP-1320, Omron Healthcare Co., Japan). The first measurement was performed after the participant rested for 15 min in a sitting position, then two additional measurements were performed 3 minutes apart. Results of the systolic and diastolic pressures were expressed in millimeter of mercury (mmHg).

#### Skin pigmentation measurements

Skin pigmentation was measured on the inner arm between biceps and triceps using the DSM III Skin colorimeter (Cortex technology, Denmark). Four measurement replicates were performed for each participant. Colors were quantified using the L*, a*, and b* parameters defined by the International commission on illumination (CIE, Vienna, Austria), where L* measures the degree of brightness or whiteness, a* the degree of redness, and b* the yellowness. Skin erythema ‘E’ and melanin content ‘M’ were also measured (57, 58).

### Biological sample collection and biobanking

#### Blood

A capillary whole blood sample was collected at the finger tip of each fasting participant using microtainer contact-activated lancets (Becton Dickinson, USA), and was immediately used for the measurement of blood glucose concentration (mg/dL) with the Freestyle Optium Neo H point-of-care device (Abbott Diabetes Care, France).

Blood samples were also collected by venipuncture in four 5 mL serum separation tubes, and in two 5 mL EDTA tubes (Becton Dickinson, USA). Serum separation tubes and EDTA tubes were maintained at 4 °C until centrifugation during 15 minutes at 3,500 and 2,000 rpm, respectively. Part of the serum and plasma samples were used by the clinical laboratory of ILM to measure several hematological and biochemical parameters as described in Table 2. The remaining samples of serum, plasma and red blood cell pellets were aliquoted in several tubes and stored at -20 °C until processing as detailed in Table 3.

**Table 2.** List of hematological and biochemical parameters analyzed for the MATAEA project.

| Group | Measured parameter | Units | Test | Analyzer |
| --- | --- | --- | --- | --- |
| **Complete blood count** | Hemoglobin | g/dL | Spectrophotometry | XT-4000 or XN-1000, Sysmex (France) |
|  | Hematocrit | % | Electrical impedance | XT-4000 or XN-1000, Sysmex (France) |
|  | Leucocytes count | G/L | Fluorocytometry | XT-4000 or XN-1000, Sysmex (France) |
|  | Neutrophils count | G/L or % | Fluorocytometry | XT-4000 or XN-1000, Sysmex (France) |
|  | Eosinophils count | G/L or % | Fluorocytometry | XT-4000 or XN-1000, Sysmex (France) |
|  | Lymphocytes count | G/L or % | Fluorocytometry | XT-4000 or XN-1000, Sysmex (France) |
|  | Monocytes count | G/L or % | Fluorocytometry | XT-4000 or XN-1000, Sysmex (France) |
|  | Platelets count | G/L | Electrical impedance | XT-4000 or XN-1000, Sysmex (France) |
|  | Mean platelet volume | fL | Electrical impedance | XT-4000 or XN-1000, Sysmex (France) |
|  | Red blood cells count | T/L | Electrical impedance | XT-4000 or XN-1000, Sysmex (France) |
|  | Red blood cell distribution width | - | Electrical impedance | XT-4000 or XN-1000, Sysmex (France) |
|  | Mean corpuscular volume | fL | Electrical impedance | XT-4000 or XN-1000, Sysmex (France) |
|  | Mean corpuscular hemoglobin | pg/cell | Spectrophotometry | XT-4000 or XN-1000, Sysmex (France) |
|  | Mean corpuscular hemoglobin concentration | g/dL | Spectrophotometry | XT-4000 or XN-1000, Sysmex (France) |
| **Serum aspect** | Serum aspect | - | Visual assessment | - |
|  | Serum index | UI | Spectrophotometry | Cobas c501, Roche (Switzerland) |
| **Diabetes** | Glycated hemoglobin A1c | % | Capillary eletrophoresis | Capillarys 2 Flex Piercing, SEBIA (France) |
|  | Fasting blood glucose level | mg/dL | Fingertip lancing technique | Optium Neo H, Abbott Diabetes Care (France) |
| **Lipids** | Total cholesterol | mmol/l | Enzymatic colorimetric assay | Cobas c501, Roche (Switzerland) |
|  | HDL-cholesterol | mmol/l | Homogenous enzymatic colorimetric assay | Cobas c501, Roche (Switzerland) |
|  | LDL-cholesterol | mmol/l | Friedewald formula (60) | Cobas c501, Roche (Switzerland) |
|  | Triglycerides | mmol/l | Enzymatic colorimetric assay | Cobas c501, Roche (Switzerland) |
| **Kidney function** | Creatinine | μmol/L | Enzymatic colorimetric assay | Cobas c501, Roche (Switzerland) |
|  | Glomerular filtration rate | mL/min/1.73 m<sup>2</sup> | Chronic Kidney Disease Epidemiology Collaboration (CKD-EPI) formula (61) | Cobas c501, Roche (Switzerland) |
| **Liver and inflammation** | Aspartate aminotransferase | UI/L | Enzymatic colorimetric assay | Cobas c501, Roche (Switzerland) |
|  | Alanine aminotransferase | UI/L | Enzymatic colorimetric assay | Cobas c501, Roche (Switzerland) |
|  | Gamma glutamyl transferase | UI/L | Enzymatic colorimetric assay | Cobas c501, Roche (Switzerland) |
|  | Total bilirubin | μmol/L | Photometry assay | Cobas c501, Roche (Switzerland) |
|  | C-reactive protein | mg/L | Particle-enhanced immunoturbidimetric assay | Cobas c501, Roche (Switzerland) |

**Table 3.** List of laboratory analyses planned in the MATAEA project.

| Sample type | Analysis | Laboratory |
| --- | --- | --- |
| Serum and plasma | Hematological and biochemical analyses | LABM, ILM, Tahiti, French Polynesia |
| Serum | Arbovirus serology | LIV, ILM, Tahiti, French Polynesia |
| Serum | Mosquito saliva serology | LEM, ILM, Tahiti, French Polynesia |
| Serum | Hantavirus serology | EIRU, IP, Paris, France |
| Serum | VirScan (62) | HEGU, IP, Paris, France |
| Plasma | Hepatitis B and C serologies | CHPF, Tahiti, French Polynesia |
| Plasma | Pesticides quantitation | INSPQ, Montreal, Canada |
| Red blood cells pellet | Heavy metals quantitation | INSPQ, Montreal, Canada |
| Red blood cells pellet, saliva | Genetic analysis | HEGU, IP, Paris, France |
| Saliva | Oral microbiome analysis | HEGU, IP, Paris, France |
| Stool | Intestinal microbiome analysis | HEGU, IP, Paris, France |

LABM: Clinical Laboratory; ILM: Institut Louis Malardé; LIV: Laboratory of research on emerging viral diseases; LEM: Laboratory of research in medical entomology; IP: Institut Pasteur; EIRU: Environment and Infectious Risk Unit ; HEGU : Human Evolutionary Genetics Unit ; CHPF: Centre Hospitalier de la Polynésie française; INSPQ: Institut National de Santé Publique du Québec.

#### Saliva

A 2 mL sample of saliva was self-collected by each participant in an Oragene DNA tube pre-filled with 2 mL of stabilization liquid following the manufacturer’s protocol (DNA Genotek, Canada). After gentle mix, the sample tube was stored at 4 °C until processing (Table 3).

#### Stool

Stool samples were self-collected by participants in tubes pre-filled with 8 mL of DNA stabilization buffer following the manufacturer’s instructions (Canvax Biotech, Spain), and stored in a fridge within 24 hours before the appointment with the nurse. Then, stool samples were stored at 4 °C until shipment to ILM. Upon reception at ILM, stool samples were stored at -20 °C until processing (Table 3).

### Data management

The data flow and processing of biological samples are detailed in Figure 3.

**Figure 3.**
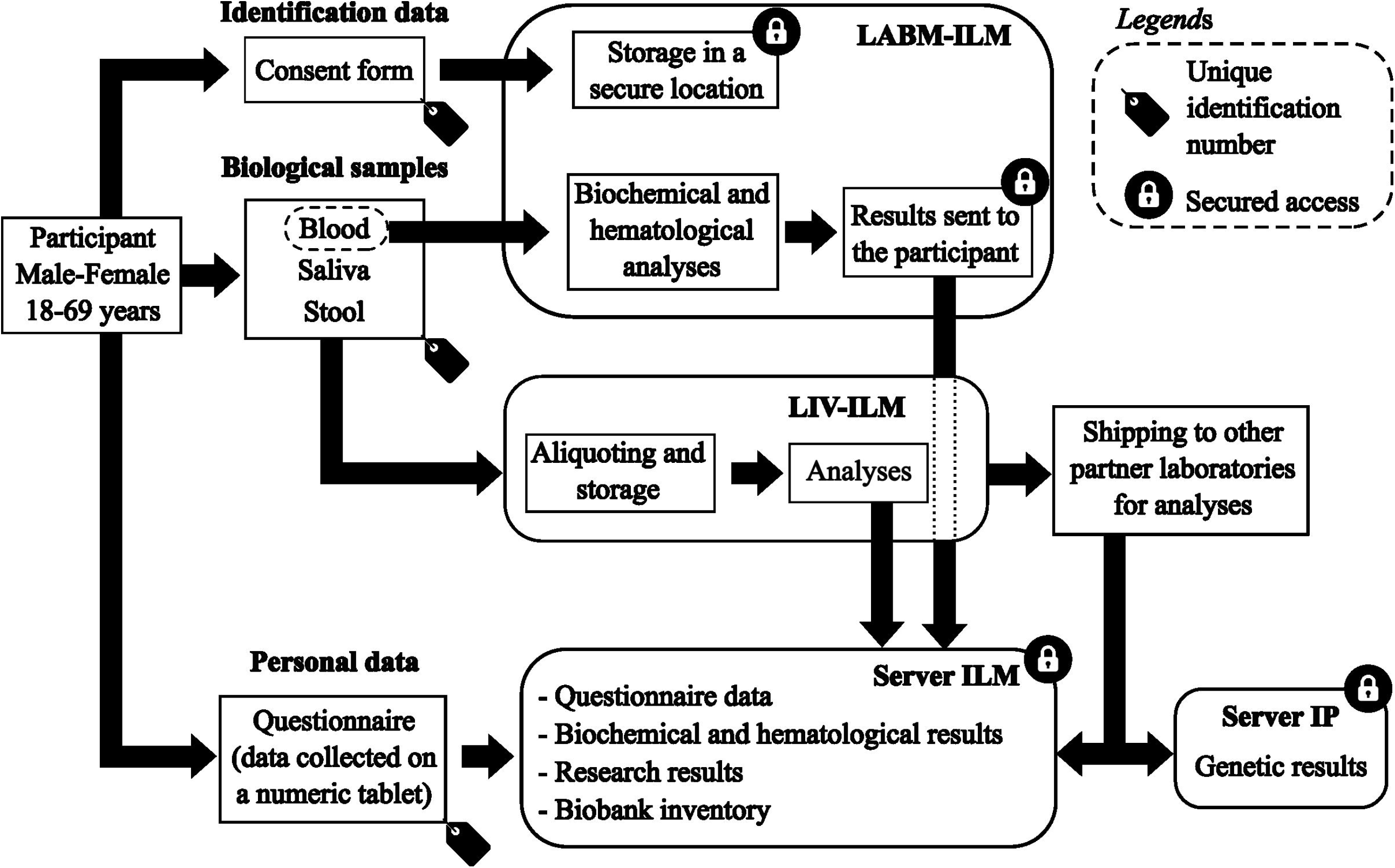
Data flow and biological samples processing for the MATAEA project. LABM: Clinical Laboratory; ILM: Institut Louis Malardé; LIV: Laboratory of research on emerging viral diseases; IP: Institut Pasteur.

Each participant was assigned a unique identification number to anonymize the information collected in the CRF and biological samples. Personal identifying information such as last name, first name, and phone number only appeared on the consent form, which has been kept in a secure location accessible only to the biologists of the clinical laboratory of ILM.

Data collected in the CRF were transferred from electronic tablets to a password-protected ODK Aggregate server (Version 2.0.5, Get ODK Inc., USA). Then, these data were uploaded to a password-protected Redcap server (Version 9.5.11, Vanderbilt University, USA) used to aggregate all data from each participant. Results of hematological and biochemical analyses performed by the clinical laboratory of ILM were also uploaded to the Redcap server, and linked to questionnaire data using the unique identification number of each participant. The results of the analyses that will be performed later from the biological samples of the participants will also be uploaded to the Redcap server. All collaborators of the MATAEA project were given a personal and unique login and password to access data in the Redcap server.

The staff of ILM received training on ethical procedures and measures to protect confidentiality needed, and was required to sign a confidentiality agreement form. The Redcap database was validated according to the guidelines of the French National Commission of Data Protection and Individual Liberties, allowing collaboration with external researchers and collaborators.

### Data Analysis

According to the data retrieved from the participants, indicators will be defined to identify subjects suffering from, or having a history, of any pathology targeted by the study.

For continuous data, summary statistics will include the arithmetic mean with 95% confidence interval (CI) or standard deviation (SD), or median with inter quartile range. For categorical data, frequency counts, percentages, and 95% CIs will be presented. The characteristics of the subjects will be compared according to sex, age category, and geographical area using the chi-square test for the categorical variables, and using the analysis of variance (ANOVA) for the continuous variables.

Non-standardized and standardized (weighted) results will be provided. Weights will be estimated by archipelago (except the Society archipelago for which the Windward Islands and Leeward Islands will be treated separately) and will correspond to the inverse of the probability for each subject to be selected, where the probability in a given archipelago is the number of subjects in each of the 6 age/sex strata divided by the total number population in that strata.

Risk factors associated with each pathology (if the prevalence is high enough to justify a statistical analysis) will be identified using a logistic regression model. The factors investigated will include age, sex, and all other relevant factors. These analyses will account for the sampling plan through consideration of survey weights.

### Return of individual results to participants

Results of physical measurements performed by a nurse were provided to each participant during the appointment. If any abnormal result was detected, the nurse advised the participant to consult a doctor immediately.

Results of hematological and biochemical analyses performed by the clinical laboratory of ILM from blood samples of each participant were available on a secured server. Participants could retrieve these results online using individual login valid for seven days, or anytime at ILM providing proof of identity. If any abnormal result was detected, the participant or the participant’s attending doctor (depending on the participant’s choice as indicated in the informed consent) was notified by a biologist of the clinical laboratory of ILM.

Participants will not be informed of individual research results that will be obtained by the specialized laboratories involved in the MATAEA project. However, if any research result suggests an underlying pathology, the participant or the participant’s attending doctor (depending on the participant’s choice as indicated in the informed consent) will be notified by a biologist of the clinical laboratory of ILM to carry out additional investigations.

## Data Availability

All data produced in the present study are available upon reasonable request to the authors

## STRENGHS AND LIMITATIONS

The MATAEA project is the first cross-sectional survey designed to investigate a broad range of health issues in the five archipelagoes of French Polynesia. The high number of participants and their wide geographical distribution guarantee a good representativeness of the population of French Polynesia. The large amount of data obtained for each participant from the CRF, physical measurements and biological analyses will make it possible to update the information on the prevalence of numerous NCDs and CDs known as major public health concerns in French Polynesia, and possibly identify specific indicators and/or risk factors for these diseases. The study will also provide new epidemiological information on poorly studied or unknown pathologies. Finally, the MATAEA project will generate a high amount of genetic data for a population that has been so far poorly represented in human genetic studies.

Nevertheless, the MATAEA project has some limitations that should be acknowledged. First, the population aged under 18 years was not included to the survey, which might have an impact on the estimate of the prevalence of certain pathologies. Second, participants were recruited in only 18 of the 75 inhabited islands while differences may exist between the inhabitants of distinct islands within the same archipelago. Third, there might be an underrepresentation of hard-to-reach subjects, such as heavy drinkers or socially excluded persons, which might be more prone to some pathologies. Fourth, the number of participants recruited for the study may be not sufficient to investigate rare pathologies. Fifth, the cross-sectional nature of the survey does not allow to examine the temporal relationship between exposures and diseases under study. Finally, the sampling period has been interrupted three times because of the COVID-19 pandemic and more than two years elapsed between the first and last inclusion (Figure 2).

## COMMUNITY ENGAGEMENT

The main objectives of the MATAEA project have already been presented to the Health and Research authorities of French Polynesia as well as, in several occasions, to the general French Polynesian populations, in particular in the form of conferences principally conducted by Van-Mai Cao-Lormeau and Lluis Quintana-Murci. Furthermore, a video presenting the MATAEA project is already accessible online (https://youtube.com/playlist?list=PLVLsoPZrkK4ipioiTOhGh0z05XZSHPC6H). All the results generated in the framework of the MATAEA project will be also made available to the population of French Polynesia through various communication channels. The results will be presented in an accessible way for the public, through conferences, seminars, and roundtables, and will be sent to the mayors of the islands that participated in the study to be relayed to the community. Finally, the public will be informed of the results published in scientific journals through popularized summaries translated into both French and Tahitian.

## ETHICS STATEMENT

This study involving human participants was approved by the *French Comité de protection des personnes* (CPP - OUEST III no. 19.08.60 / SI CNRIPH 19.07.02.38421) and the *Comité d’Ethique de la Polynésie française* (Avis no. 80 CEPF-03/09/2019). Written informed consent was obtained from all participants. People who were unable to express their consent or answer the questionnaire were excluded from the study. Data collection, storage, transfer and process were executed according to the European general data protection regulation procedures.

## AUTHOR CONTRIBUTIONS

AF, LQM, AS, and VMCL designed and conceptualized the project; AF, LQM, EP, AS, JV, NP, JT, SO, SL, HB, PB, CG, ES, BC and PA participated in the design of the study protocol; IT, MA, and VMCL wrote the study protocol; SFP and NJ managed the ethical aspects of the project; NP and JT carried out the draw of participants; IT, MA, and VMCL supervised and coordinated the project; IT, EP, AJ, MR, VM, and KC performed the data management; CH and AB processed biological samples; GRL, DL and EP designed genetic analyses; VMCL acquired funding for the project; IT and MA drafted the manuscript. All authors read and approved the submitted version of the manuscript.

## ACKNOWLEDGEMENTS

We thank the staff of all the laboratories that contributed to the realization of this study. We also acknowledge the municipal staff and the guides in the islands selected for the study for their support in recruiting participants. Finally, we especially thank all the individuals who willingly agreed to participate in this study

