## Supplemental Data for "Unravelling the determinants of human health in French Polynesia: the MATAEA project"

***Supplementary Material***

**Supplementary Data 1. Complete case report form for the MATAEA project.**

| **Survey information** | | | |
| --- | --- | --- | --- |
|  | **Question** | **Answer** | |
|  | **Unique identification number** | **꙱꙱꙱꙱꙱꙱꙱꙱꙱** | |
| #1 | Household ID | _ _ _ _ | |
| #2 | City ID | _ _ | |
| #3 | City name | _ _ _ _ _ _ | |
| #4 | Interviewer ID | _ _ | |
| #5 | Date of interview | _ _ _ _ _ _ _ _  Day Month Year | |
| #6 | Interview Language | French  Tahitian | 1  2 |
| #7 | Time of interview (24 hour clock) | _ _ : _ _  hr min | |
| #8 | Consent has been read and obtained | Yes  No | 1  2 **if NO, END** |

| **Demographic Information** | | | |
| --- | --- | --- | --- |
|  | **Question** | **Answer** | |
| #9 | Sex | Male | 1 |
|  |  | Female | 2 |
| #10 | What is your date of birth?  *Don't Know 77 77 7777* | _ _ _ _ _ _ _ _  *Day Month Year*  *If Known, Go to #12* | |
| #11 | How old are you? | Years | _ _ |
| #12 | In total, how many years have you spent at school and in full-time study (excluding pre-school)? | Years | _ _ |
| #13 | What is the highest level of education you have completed? | No formal schooling  Less than primary school  Primary school completed  Secondary school completed  High school completed  College/University completed  Post graduate degree  Don’t want to answer | 1  2  3  4  5  6  7  88 |
| #14 | Where were you born? | *French Polynesia*  *France (except French Polynesia)*  Other  Don’t know  Don’t want to answer | *987*  *1*  *2*  *77*  *88* |
| #15 | What is your ethnic group background? | Polynesian  Caucasian  Asian  Mixed  Other  Don’t know  Don’t want to answer | 1  2  3  4  5  77  88 |
|  |  | Other (please specify): | …………………… |
| #16 | What is your marital status? | Never married  Currently married  Separated  Divorced  Widowed  Cohabitating  Don’t want to answer | 1  2  3  4  5  6  88 |
| #17 | Which of the following best describes your main work status over the past 12 months?  *(USE SHOWCARD)* | Government employee  Non-government employee  Self-employed  Non-paid  Student  Homemaker  Retired  Unemployed (able to work)  Unemployed (unable to work)  Don’t want to answer | 1  2  3  4  5  6  7  8  9  88 |
| #18 | How many people older than 18 years, including yourself, live in your household? | Number of people | _ _ |
| #19 | How many people, including yourself, live in your household? | Number of people | _ _ |
| **Family history** | | | |
|  | **Question** | **Answer** | |
| #20 | Place of birth | In French Polynesia  Outside of French Polynesia | 1  2 |
| #21 | If you were not born in French Polynesia, for how many years have you lived in French Polynesia? | Years  Don’t want to answer | _ _  88 |
| #22 | Place of birth of both parents  *Indicate an island (if born in one of the French overseas territories) or a country (if born in France or another country)*  *Don’t know 77*  *Don’t want to answer 88* | Mother  Father | ………………..  ………………… |
| #23 | Birth place of the four grandparents  *Indicate an island (if born in one of the French overseas territories) or a country (if born in France or another country)*  *Don’t know 77*  *Don’t want to answer 88* | Paternal grandfather  Paternal grandmother  Maternal grandfather  Maternal grandmother | ………………..  …………………  …………………  …………………. |
| #24 | Native language | French  Tahitian  English  Spanish  Other | 1  2  3  4  5 |
|  |  | Other (please specify): | …………………… |

| **Tobacco use** | | | | | | | | | |
| --- | --- | --- | --- | --- | --- | --- | --- | --- | --- |
|  | **Question** | **Answer** | | | | | | | |
| #25 | Do you currently smoke any tobacco products, such as cigarettes, cigars or pipes?  *(USE SHOWCARD)* | Yes  No | | | | 1  2 *If No, go to #32* | | | |
| #26 | Do you currently smoke tobacco products daily? | Yes  No | | | | 1  2 | | | |
| #27 | How old were you when you first started smoking? | Age (Years)  Don’t know | _ _ *If Known, go to #29*  77 | | | | | | |
| #28 | Do you remember how long ago it was?  *(RECORD ONLY 1, NOT ALL 3)*  *Don’t know 77* | In Years | _ _ *If Known, go to #29* | | | | | | |
|  |  | OR in Months | _ _ *If Known, go to #29* | | | | | | |
|  |  | OR in Weeks | _ _ | | | | | | |
| #29 | On average, how many of the following products do you smoke each day/week?  *(IF LESS THAN DAILY, RECORD WEEKLY)*  *(RECORD FOR EACH TYPE, USE SHOWCARD)*  *Don’t know 7777* |  | | | | DAILY**↓** WEEKLY↓ | | | |
|  |  | Manufactured cigarettes | | | | _ _ _ _ _ _ _ _ | | | |
|  |  | Hand-rolled cigarettes | | | | _ _ _ _ _ _ _ _ | | | |
|  |  | Other | | | | _ _ _ _ _ _ _ _ | | | |
|  |  | Other (please specify): | | | | …………………… | | | |
| #30 | During the past 12 months, have you tried to stop smoking? | Yes  No | | | | 1  2 | | | |
| #31 | During any visit to a doctor or other health worker in the past 12 months, were you advised to quit smoking tobacco? | Yes  No  No visit during the past 12 months | | | | | | 1  2  3 | |
| #32 | In the past, did you ever smoke any tobacco products? *(USE SHOWCARD)* | Yes  No | | | | 1 *If Yes, go to #33*  2 *If No, go to #34* | | | |
| #33 | In the past, did you ever smoke daily? | Yes  No | | | | 1  2 | | | |
| #34 | During the past 30 days, did someone smoke in your home? | Yes  No | | | | 1  2 | | | |
| #35 | During the past 30 days, did someone smoke in closed areas in your workplace (in the building, in a work area or a specific office)? | Yes  No  Don't work in a closed area | | | | 1  2  3 | | | |
| **Cannabis use** | | | | | | | | | |
|  | **Question** | **Answer** | | | | | | | |
| #36 | Have you ever smoked cannabis? | Yes  No  Don’t know  Don’t want to answer | | 1  2 *If No or Don’t want to answer, go to #42*  77  88 | | | | | |
| #37 | How old were you when you first started smoking cannabis? (USE SHOWCARD) | Don’t know  Don’t want to answer | | | _ _  77  88 | | | | |
| #38 | In the past 12 months, did you smoke cannabis? | Yes | | | 1 | | | | |
|  |  | No | | | 2 *If No or Don’t want to answer, go to #42* | | | | |
|  |  | Don’t know | | | 77 | | | | |
|  |  | Don’t want to answer | | | 88 | | | | |
| #39 | In the past 12 months, how often did you smoke cannabis? | 5-7 days per week  2-4 days per week  1 day per week  1-3 days per month  Less than once a month  Don’t know  Don’t want to answer | | | 1  2  3  4  5  77  88 | | | | |
| #40 | At what age did you start smoking cannabis at least once a week?  *(USE SHOWCARD)* | Never  Don’t know  Don’t want to answer | | | _ _  99  77  88 | | | | |
| #41 | When smoking cannabis, do you consume alcohol at the same time? | Never  Sometimes  Often  Always  Don’t know  Don’t want to answer | | | 1  2  3  4  77  88 | | | | |
| **Alcohol Consumption** | | | | | | | | | |
|  | **Question** | **Answer** | | | | | | | |
| #42 | Have you ever consumed an alcoholic beverage such as beer, wine, liqueur, cider, whiskey, rum or champagne? *(USE SHOWCARD)* | Yes  No | | | | 1  2 *If No, go to #53* | | | |
| #43 | Have you consumed any alcohol within the past 12 months? | Yes  No | | | | 1 *If Yes, go to #45*  2 | | | |
| #44 | Have you stopped drinking due to health reasons, such as a negative impact on your health or on the advice of your doctor or other health worker? | Yes  No | | | | 1 *If Yes, go to #52*  2 *If No, go to #52* | | | |
| #45 | During the past 12 months, how frequently have you had at least one standard alcoholic drink?  *(USE SHOWCARD)* | Daily  5-6 days per week  3-4 days per week  1-2 days per week  1-3 days per month  Less than once a month  Never | | | | 1  2  3  4  5  6  7 | | | |
| #46 | Have you consumed any alcohol within the past 30 days? | Yes  No | | | | 1  2 *If No, go to #52* | | | |
| #47 | During the past 30 days, on how many occasions did you have at least one standard alcoholic drink? | Number  Don’t know | | | | _ _ *If zero, go to #52*  *77* | | | |
| #48 | During the past 30 days, when you drank alcohol, how many standard drinks on average did you have during one drinking occasion?  *(USE SHOWCARD)* | Number  Don’t know | | | | _ _  77 | | | |
| #49 | During the past 30 days, what was the largest number of standard drinks you had on a single occasion, counting all types of alcoholic drinks together? | Largest number  Don’t know | | | | _ _  77 | | | |
| #50 | During the past 30 days, how many times did you have six or more standard drinks in a single drinking occasion? | Number of times Don’t know | | | | _ _  77 | | | |
| #51 | During each of the past 7 days, how many standard drinks did you have each day?  *(USE SHOWCARD)*  *Don’t know 77* | Monday | | | | _ _ | | | |
|  |  | Tuesday | | | | _ _ | | | |
|  |  | Wednesday | | | | _ _ | | | |
|  |  | Thursday | | | | _ _ | | | |
|  |  | Friday | | | | _ _ | | | |
|  |  | Saturday | | | | _ _ | | | |
|  |  | Sunday | | | | _ _ | | | |
| #52 | During the past 12 months, have you had family problems or problems with your partner due to your drinking? | Yes, more than monthly  Yes, monthly  Yes, several times but less than monthly  Yes, once or twice  No | | | | | | | 1  2  3  4  5 |
| **Diet** | | | | | | | | | |
|  | **Question** | **Answer** | | | | | | | |
| #53 | In a typical week, on how many days do you eat fruit? *(USE SHOWCARD)* |  | | | |  | | | |
|  |  | Number of days | | | | _ _ | | | |
|  |  | Don’t know | | | | 77 | | | |
| #54 | How many servings of fruit do you eat on one of those days? (*USE SHOWCARD)* | Number of servings  Don’t know | | | | _ _  77 | | | |
| #55 | What is the main reason you do not consume more fruit?  *(SELECT ONLY ONE)* | My consumption seems sufficient | | | | 1 | | | |
|  |  | The price | | | | 2 | | | |
|  |  | Difficulties of supply | | | | 3 | | | |
|  |  | Binding preparation | | | | 4 | | | |
|  |  | Presence of pesticides | | | | 5 | | | |
|  |  | No fruit in the garden | | | | 6 | | | |
|  |  | I don’t like fruits | | | | 7 | | | |
|  |  | Other | | | | 8 | | | |
|  |  | Other (please, specifiy): | | | | …………………… | | | |
| #56 | In a typical week, on how many days do you eat vegetables? *(USE SHOWCARD)* |  | | | |  | | | |
|  |  | Number of days | | | | _ _ | | | |
|  |  | Don’t know | | | | 77 | | | |
| #57 | How many servings of vegetables do you eat on one of those days? *(USE SHOWCARD)* | Number of servings  Don’t know | | | | _ _  77 | | | |
| #58 | What is the main reason why you don't eat more vegetables?  *(SELECT ONLY ONE)* | My consumption seems sufficient | | | | 1 | | | |
|  |  | The price | | | | 2 | | | |
|  |  | Difficulties of supply | | | | 3 | | | |
|  |  | Binding preparation | | | | 4 | | | |
|  |  | Presence of pesticides | | | | 5 | | | |
|  |  | No vegetable patch | | | | 6 | | | |
|  |  | I don’t like vegetables | | | | 7 | | | |
|  |  | Other | | | | 8 | | | |
|  |  | Other (please, specify): | | | | …………………… | | | |
| #59 | In a typical week, on how many days do you eat meat? *(USE SHOWCARD)* | Number of days  Don’t know | | | | _ _  77 | | | |
| #60 | How many servings of meat do you eat on one of those days? *(USE SHOWCARD)* | Number of servings  Don’t know | | | | _ _  77 | | | |
| #61 | What is the main reason why you don't eat more meat?  *(SELECT ONLY ONE)* | The price  Difficulties of supply Binding preparation  Presence of antibiotics  I don’t like meat  Other  Don’t know  Don’t want to answer  Other (please, specify) | | | | 1  2  3  4  5  6  77  88  …………………… | | | |
| #62 | In a typical week, on how many days do you eat fish?  *(USE SHOWCARD)* | Number of days  Don’t know | | | | _ _  77 | | | |
| #63 | How many servings of fish do you eat on one of those days? *(USE SHOWCARD)* | Number of servings  Don’t know | | | | _ _  77 | | | |
| #64 | What is the main reason why you don't eat more fish?  *(SELECT ONLY ONE)* | The price  Difficulties of supply  Binding preparation  Presence of pesticides  I don’t like fish  Other  Don’t know  Don’t want to answer  Other (please, specify): | | | | 1  2  3  4  5  6  77  88  …………………… | | | |
| #65 | How often do you add salt or a salty sauce such as soy sauce to your food right before you eat it or as you are eating it?  *(SELECT ONLY ONE)*  *(USE SHOWCARD)* | Always  Often  Sometimes  Rarely  Never  Don’t know | | | | 1  2  3  4  5  77 | | | |
| #66 | How often is salt, salty seasoning or a salty sauce added in cooking or preparing foods in your household? | Always  Often  Sometimes  Rarely  Never  Don’t know | | | | 1  2  3  4  5  77 | | | |
| #67 | How often do you eat processed food high in salt? By processed food high in salt, I mean foods that have been altered from their natural state, such as packaged salty snacks, canned salty food including pickles and preserves, salty food prepared at a fast food restaurant, cheese, bacon and processed meat*. (USE SHOWCARD)* | Always  Often  Sometimes  Rarely  Never  Don’t know | | | | 1  2  3  4  5  77 | | | |
| #68 | How much salt or salty sauce do you think you consume? | Far too much  Too much  Just the right amount  Too little  Far too little  Don’t know | | | | 1  2  3  4  5  77 | | | |
| #69 | Do you think that too much salt or salty sauce in your diet could cause a health problem? | Yes  No  Don’t know | | | | 1  2  77 | | | |
| Do you do any of the following on a regular basis to control your salt intake? *(RECORD FOR EACH)* | | | | | | | | | |
| #70 | Limit consumption of processed foods | Yes  No | | | | 1  2 | | | |
| #71 | Look at the salt or sodium content on food labels | Yes  No | | | | 1  2 | | | |
| #72 | Use spices other than salt when cooking | Yes  No | | | | 1  2 | | | |
| #73 | Avoid eating foods prepared outside of a home | Yes  No | | | | 1  2 | | | |
| #74 | In the past 30 days, how often have you consumed sugary drinks such as soda, coke, syrups or fruit juice?  *(USE SHOWCARD)* | 5 to 7 days a week  2 to 4 days a week  1 day a week  1 to 3 days per month  Less than one day per month  Don’t know  Don’t want to answer | | | | 1  2  3  4  5  77  88 *If Don’t want to answer, go to #77* | | | |
| #75 | How many glasses do you have on average on one of these days?  *(USE SHOWCARD)* | Number of drinks  Don’t know  Don’t want to answer | | | | _ _  77  88 | | | |
| #76 | When do you usually drink these sugary drinks? By meal, we mean breakfast, lunch and dinner  *(USE SHOWCARD)* | During meals  Between meals  At meals and between meals Don’t know  Don’t want to answer | | | | 1  2  3  77  88 | | | |
| #77 | How often do you eat foods such as potato chips, spring rolls, ice cream, pastries, sweet, cakes, chocolate, fast food, etc.  *(USE SHOWCARD)* | 5 to 7 days a week  2 to 4 days a week  1 day a week  1 to 3 days per month  Less than one day per month  Don’t know  Don’t want to answer | | | | 1  2  3  4  5  77  88 | | | |
| **Physical activities** | | | | | | | | | |
|  | **Question** | **Answer** | | | | | | | |
| #78 | Does your work involve vigorous-intensity activity that causes large increases in breathing or heart rate like *[carrying or lifting* *heavy loads, digging or construction work]* for at least 10 minutes continuously?  *(USE SHOWCARD)* | Yes  No | | | | 1  2 *If No, go to #81* | | | |
| #79 | In a typical week, on how many days do you do vigorous-intensity activities as part of your work? | Number of days | | | | _ | | | |
| #80 | How much time do you spend doing vigorous-intensity activities at work on a typical day? | Hours : minutes | | | | _ _: _ _ | | | |
| #81 | Does your work involve moderate-intensity activity that causes small increases in breathing or heart rate such as brisk walking *[or carrying light loads]* for at least 10 minutes continuously?  *(USE SHOWCARD)* | Yes  No | | | | 1  2 *If No, go to #84* | | | |
| #82 | In a typical week, on how many days do you do moderate-intensity activities as part of your work? | Number of days | | | | _ | | | |
| #83 | How much time do you spend doing moderate-intensity activities at work on a typical day? | Hours : minutes | | | | _ _ : _ _ | | | |
| #84 | Do you walk or use a bicycle *(pedal cycle)* for at least 10 minutes continuously to get to and from places? | Yes  No | | | | 1  2 *If No, go to #87* | | | |
| #85 | In a typical week, on how many days do you walk or bicycle for at least 10 minutes continuously to get to and from places? | Number of days | | | | _ _ | | | |
| #86 | How much time do you spend walking or bicycling for travel on a typical day? | Hours : minutes | | | | _ _ : _ _ | | | |
| #87 | Do you do any vigorous-intensity sports, fitness or recreational *(leisure)* activities that cause large increases in breathing or heart rate like *[running or football]* for at least 10 minutes continuously?  *(USE SHOWCARD)* | Yes  No | | | | 1  2 *If No, go to #90* | | | |
| #88 | In a typical week, on how many days do you do vigorous-intensity sports, fitness or recreational *(leisure)* activities? | Number of days | | | | _ | | | |
| #89 | How much time do you spend doing vigorous-intensity sports, fitness or recreational activities on a typical day? | Hours : minutes | | | | _ _ : _ _ | | | |
| #90 | Do you do any moderate-intensity sports, fitness or recreational *(leisure)* activities that cause a small increase in breathing or heart rate such as brisk walking*, [cycling, swimming and volleyball]* for at least 10 minutes continuously?  *(USE SHOWCARD)* | Yes  No | | | | 1  2 | | | |
| #91 | In a typical week, on how many days do you do moderate-intensity sports, fitness or recreational *(leisure)* activities? | Number of days | | | | _ _ | | | |
| #92 | How much time do you spend doing moderate-intensity sports, fitness or recreational *(leisure)* activities on a typical day? | Hours : minutes | | | | _ _ : _ _ | | | |
| #93 | How much time do you usually spend sitting or reclining on a typical day? | Hours : minutes | | | | _ _ : _ _ | | | |
|  | **Question** | **Answer** | | | | | | | |
| #94 | In general, would you say your health is: | Excellent  Very good  Good  Fair  Poor | | | | 1  2  3  4  5 | | | |
| The following items are about activities you might do during a typical day. Does your health now limit you in these activities? If so, how much? | | | | | | | | | |
| #95 | Moderate activities, such as moving a table, vaccuming or bowling | Yes, limited a lot  Yes, limited a little  No, not limited at all | | | | 1  2  3 | | | |
| #96 | Climbing several flights of stairs | Yes, limited a lot  Yes, limited a little  No, not limited at all | | | | 1  2  3 | | | |
| During the past 4 weeks, have you had any of the following problems with your work or other regular daily activities as a result of any emotional problems (such as feeling depressed or anxious)? | | | | | | | | | |
| #97 | Accomplished less than you would like | All of the Time  Most of the Time  Some of the Time  A Little of the Time  None of the Time | | | | 1  2  3  4  5 | | | |
| #98 | Didn’t do work or other activities as carefully as usual | All of the Time  Most of the Time  Some of the Time  A Little of the Time  None of the Time | | | | 1  2  3  4  5 | | | |
| During the past 4 weeks, have you had any of the following problems with your work or other regular daily activities as a result of your physical health? | | | | | | | | | |
| #99 | Accomplished less than you would like | All of the Time  Most of the Time  Some of the Time  A Little of the Time  None of the Time | | | | | 1  2  3  4  5 | | |
| #100 | Were limited in the kind of work or other activities | All of the Time  Most of the Time  Some of the Time  A Little of the Time  None of the Time | | | | | 1  2  3  4  5 | | |
| #101 | During the PAST 4 WEEKS, how much did PAIN interfere with your normal work (including both work outside the home and housework)? | Not at all  A little bit  Moderately  Quite a bit  Extremely | | | | | 1  2  3  4  5 | | |
| The following questions are about how you felt during the past 4 weeks. For each question, indicate the answer that you think is most appropriate: | | | | | | | | | |
| #102 | Have you felt calm and peaceful? | All of the Time  Most of the Time  Some of the Time  A Little of the Time  None of the Time | | | | 1  2  3  4  5 | | | |
| #103 | Did you have a lot of energy? | All of the Time  Most of the Time  Some of the Time  A Little of the Time  None of the Time | | | | 1  2  3  4  5 | | | |
| #104 | Have you felt downhearted and blue? | All of the Time  Most of the Time  Some of the Time  A Little of the Time  None of the Time | | | | 1  2  3  4  5 | | | |
| #105 | During the past 4 weeks, how much of the time has your physical health or emotional problems interfered with your social activities (like visiting with friends, relatives, etc.)? | All of the Time  Most of the Time  Some of the Time  A Little of the Time  None of the Time | | | | 1  2  3  4  5 | | | |
| **History of Raised Blood Pressure** | | | | | | | | | |
|  | **Question** | **Answer** | | | | | | | |
| #106 | Have you ever had your blood pressure measured by a doctor or other health worker? | Yes  No | | | | 1  2 *If No, go to #112* | | | |
| #107 | Have you ever been told by a doctor or other health worker that you have raised blood pressure or hypertension? | Yes  No | | | | 1  2 *If No, go to #112* | | | |
| #108 | Were you first told in the past 12 months? | Yes  No | | | | 1  2 | | | |
| #109 | In the past two weeks, have you taken any drugs (medication) for raised blood pressure prescribed by a doctor or other health worker? | Yes  No | | | | 1  2 | | | |
| #110 | Have you ever seen a traditional healer for raised blood pressure or hypertension? | Yes  No | | | | 1  2 | | | |
| #111 | Are you currently taking any herbal or traditional remedy for your raised blood pressure? | Yes  No | | | | 1  2 | | | |
| **History of Diabetes** | | | | | | | | | |
|  | **Question** | **Answer** | | | | | | | |
| #112 | Have you ever had your blood sugar measured by a doctor or other health worker? | Yes  No | | | | 1  2 *If No, go to #119* | | | |
| #113 | Have you ever been told by a doctor or other health worker that you have raised blood sugar or diabetes? | Yes  No | | | | 1  2 *If No, go to #119* | | | |
| #114 | Were you first told in the past 12 months? | Yes  No | | | | 1  2 | | | |
| #115 | In the past two weeks, have you taken any drugs (medication) for diabetes prescribed by a doctor or other health worker? | Yes  No | | | | 1  2 | | | |
| #116 | Are you currently taking insulin for diabetes prescribed by a doctor or other health worker? | Yes  No | | | | 1  2 | | | |
| #117 | Have you ever seen a traditional healer for diabetes or raised blood sugar? | Yes  No | | | | 1  2 | | | |
| #118 | Are you currently taking any herbal or traditional remedy for your diabetes? | Yes  No | | | | 1  2 | | | |
| **History of Raised Total Cholesterol** | | | | | | | | | |
|  | **Question** | **Answer** | | | | | | | |
| #119 | Have you ever had your cholesterol (fat levels in your blood) measured by a doctor or other health worker? | Yes  No | | | | 1  2 *If No, go to #125* | | | |
| #120 | Have you ever been told by a doctor or other health worker that you have raised cholesterol? | Yes  No | | | | 1  2 *If No, go to #125* | | | |
| #121 | Were you first told in the past 12 months? | Yes  No | | | | 1  2 | | | |
| #122 | In the past two weeks, have you taken any oral treatment (medication) for raised total cholesterol prescribed by a doctor or other health worker? | Yes  No | | | | 1  2 | | | |
| #123 | Have you ever seen a traditional healer for raised cholesterol? | Yes  No | | | | 1  2 | | | |
| #124 | Are you currently taking any herbal or traditional remedy for your raised cholesterol? | Yes  No | | | | 1  2 | | | |
| **History of Cardiovascular Diseases** | | | | | | | | | |
|  | **Question** | **Answer** | | | | | | | |
| #125 | Have you ever had a heart attack or chest pain from heart disease (angina) or a stroke (cerebrovascular accident or incident)? | Yes  No | | | | 1  2 | | | |
| **Lifestyle Advice** | | | | | | | | | |
|  | **Question** | **Answer** | | | | | | | |
| #126 | During the past 12 months, have you visited a doctor or other health worker? | Yes  No | | | | 1  2 | | | |
| During any of your visits to a doctor or other health worker in the past 12 months, were you advised to do any of the following? (RECORD FOR EACH) | | | | | | | | | |
| #127 | Quit using tobacco or don’t start | Yes  No | | | | 1  2 | | | |
| #128 | Reduce salt in your diet | Yes  No | | | | 1  2 | | | |
| #129 | Eat at least five servings of fruit and/or vegetables each day | Yes  No | | | | 1  2 | | | |
| #130 | Reduce fat in your diet | Yes  No | | | | 1  2 | | | |
| #131 | Start or do more physical activity | Yes  No | | | | 1  2 | | | |
| #132 | Maintain a healthy body weight or lose weight | Yes  No | | | | 1  2 | | | |
| #133 | Reduce sugary beverages in your diet | Yes  No | | | | 1  2 | | | |
| **CORE (for women only): Cervical Cancer Screening** | | | | | | | | | |
| The next question asks about cervical cancer prevention. Screening tests for cervical cancer prevention can be done in different ways, including Visual Inspection with Acetic Acid/vinegar (VIA), pap smear and Human Papillomavirus (HPV) test. VIA is an inspection of the surface of the uterine cervix after acetic acid (or vinegar) has been applied to it. For both pap smear and HPV test, a doctor or nurse uses a swab to wipe from inside your vagina, take a sample and send it to a laboratory. It is even possible that you were given the swab yourself and asked to swab the inside of your vagina. The laboratory checks for abnormal cell changes if a pap smear is done, and for the HP virus if an HPV test is done. | | | | | | | | | |
|  | **Question** | **Answer** | | | | | | | |
| #134 | Have you ever had a screening test for cervical cancer, using any of these methods described above? | Yes  No  Don’t know | | | | 1  2  77 | | | |
| **Long-term illness** | | | | | | | | | |
|  | **Question** | **Answer** | | | | | | | |
| #135 | Do you have a long-term illness? | Yes  No | | | | 1  2 *If No, go to #137* | | | |
| #136 | Please, specify: | Hypertension  Diabetes  Other  Don’t know | | | | 1  2  3  77 | | | |
|  |  | If Other (please specifiy) | | | | ………… | | | |
| **Allergies** | | | | | | | | | |
|  | **Question** | **Answer** | | | | | | | |
| #137 | Have you ever been told by a doctor or other health professional that you had any kind of food/digestive allergy (dairy, gluten, eggs, peanuts, fish …) | Yes  No  Don’t know | | | | 1  2  77 | | | |
| #138 | Have you ever been told by a doctor or other health professional that you had any kind of respiratory allergy (dust, pollen, animals…) | Yes  No  Don’t know | | | | 1  2  77 | | | |
| #139 | Have you ever been told by a doctor or other health professional that you had any kind of skin allergy (dust, pollen, animals…) | Yes  No  Don’t know | | | | 1  2  77 | | | |
| **Respiratory diseases** | | | | | | | | | |
|  | **Question** | **Answer** | | | | | | | |
| #140 | Have you ever been told by a doctor or other health professional that you had asthma? | Yes  No  Don’t know  Don’t want to answer | | | | 1  2  77  88 | | | |
| #141 | In the past 12 months, have you had any wheezing or whistling in your chest? | Yes  No  Don’t know  Don’t want to answer | | | | 1  2  77  88 | | | |
| #142 | In the past 12 months, has your chest sounded wheezy during or after exercise or physical activity? | Yes  No  Don’t know  Don’t want to answer | | | | 1  2  77  88 | | | |
| #143 | In a year, do you cough on most days for at least 3 consecutive months or more? | Yes  No  Don’t know  Don’t want to answer | | | | 1  2  77  88 | | | |
| #144 | For how many years have you had this cough? | Years  Don’t know  Don’t want to answer | | | | _ _  77  88 | | | |
| #145 | In a year, do you have expectoration (bring up phlegm) on most days for at least 3 consecutive months or more? | Yes  No  Don’t know  Don’t want to answer | | | | 1  2  77  88 | | | |
| #146 | For how many years have you had trouble with phlegm? | Years  Don’t know  Don’t want to answer | | | | _ _  77  88 | | | |
| **Cancer** | | | | | | | | | |
|  | **Question** | **Answer** | | | | | | | |
| #147 | Have you ever been told by a doctor or other  health professional that you had cancer or a  malignant tumor of any kind? | Yes  No  Don’t know  Don’t want to answer | | | | 1  2  77  88 | | | |
| #148 | What kind of cancer was it?  (3 answers maximum) | Breast  Cervix  Lung  Colorectal  Prostate  Other  More than 3 types of cancer | | | | 1  2  3  4  5  6  8 | | | |
|  |  | If Other (please specify): | | | | ………… | | | |
| #149 | How old were you when this cancer was diagnosed for the first time? | Years  Don’t know  Don’t want to answer | | | | _ _  77  88 | | | |
| #150 | Whether dead or alive, has your mother / father / brother / sister or child ever had cancer or a malignant tumor? | Yes  No  Don’t know  Don’t want to answer | | | | 1  2  77  88 | | | |
| **Disabilities** | | | | | | | | | |
|  | **Question** | **Answer** | | | | | | | |
| #151 | Does your physical, mental or emotional health hinder you in your daily activities? | Yes  No  Don’t know  Don’t want to answer | | | | 1  2  77  88 | | | |
| #152 | Are you a blind or visually impaired personor have serious difficulty seeing even when wearing glasses? | Yes  No  Don’t know  Don’t want to answer | | | | 1  2  77  88 | | | |
| #153 | Are you a deaf person or have serious difficulty hearing? | Yes  No  Don’t know  Don’t want to answer | | | | 1  2  77  88 | | | |
| **Ciguatera** | | | | | | | | | |
|  | **Question** | **Answer** | | | | | | | |
| #154 | How many times have you had Ciguatera? | 0  1  2  3  4  Between 5 and 9  More then 10  Don’t know | | | | 0  1  2  3  4  5  6  77 | | | |
| #155 | When you were poisoned, did you seek medical attention? | Yes (everytime)  Yes (but not everytime)  No | | | | 1  2  3 | | | |
| #156 | If No, why didn’t you consult a medical structure?  *(several possible answers)* | Symptoms were not severe  It is useless because there is no effective treatment  I prefer to treat myself with traditional medicine  I didn't want to pay for medical consultation just for that  Other | | | | 1  2  3  4  5 | | | |
|  |  | If Other (please specify): | | | | …………………… | | | |
| #157 | If you have had several episodes of ciguatera, which one was the most severe | The first one  The last one  Neither the first nor the last | | | | 1  2  3 | | | |
| #158 | How quickly did all of your symptoms disappear (including itching, tingling, fatigue…)? | Less than a month  Between 1 and 3 months  Between 3 and 6 months  Between 6 months and a year  More than a year  Don’t know | | | | 1  2  3  4  5  77 | | | |
| #159 | Which marine products got you ill? (if several CF poisoning, list fish species, from the first to the last poisoning) | …………………… | | | | | | | |
| #160 | How was the fish/marine product obtained? | Personal / family fishing / given by a friend  Bought directly from the fisherman  Bought by the roadside  Purchased in supermarkets / stores / markets | | | | 1  2  3  4 | | | |
| #161 | Were you aware that there was a risk of being poisoned by consuming these fish? | Yes  Only for some (if several ciguatera)  No | | | | 1  2  3 | | | |
| #162 | Did you change your fish consumption as a result of your poisoning? *(several possible answers)* | Yes, I eat less or no fish at all  I eat more offshore fish  I no longer consume certain risk species  No, I haven't changed anything | | | | 1  2  3  4 | | | |
| **Biomarkers of exposure to mosquitoes** | | | | | | | | | |
|  | **Question** | **Answer** | | | | | | | |
| #163 | Is your accommodation: | A house with a garden  A house without a garden  An apartment with terrace  An apartment without terrace | | | | 1  2  3  4 | | | |
| #164 | Is your accommodation air conditioned? | Yes  No | | | | 1  2 | | | |
| #165 | Do you have access to running water in your home? | Yes  No | | | | 1  2 | | | |
| #166 | Do you get bitten by mosquitoes? | Never  Rarely  Often  Everyday | | | | 1  2  3  4 | | | |
| #167 | What means do you use to protect yourself from mosquito bites?  *(several possible answers)* | Elimination of larval breeding sites  Spraying of insecticides  Application of repellent products for the skin  Mosquito coils  Plant fires (smoke)  Use of mosquito nets  None  Mosquito rackets  Fan | | | | 1  2  3  4  5  6  7  8  9 | | | |
| **COVID-19** | | | | | | | | | |
|  | **Question** | **Answer** | | | | | | | |
| #168 | Have you ever been tested positive for covid-19? (Nasopharyngeal test performed by medical staff)  *If Yes, go to #169*  *If No, go to #177* | Yes  No  Don’t know | | | | 1  2  77 | | | |
| #169 | If yes, when did you test positive? | Date | | | | _ _ _ _ _ _ | | | |
| #170 | How long did you stay isolated? | Number of days | | | | _ _ | | | |
| #171 | *Did you have the following symptoms?*  *(several possible answers)* | Fever (≥38 °C)  Sore throat  Runny nose  Cough  Shortness of breath  Vomiting  Nausea  Diarrhea  New loss of smell or taste  Asymptomatic | | | | 1  2  3  4  5  6  7  8  9  10 | | | |
| #172 | What was your medical treatment? | None  Medical consultation  Traditional medicine | | | | 1  2  3 | | | |
| #173 | How long have you experienced symptoms? | Asymptomatic  Less than a week  1 week to 1 month  More than a month | | | | 1  2  3  4 | | | |
| #174 | Have you had any complications related to COVID-19? | Yes  No | | | | 1  2 | | | |
| #175 | What were the complications of your illness? | Need for “simple” hospitalization  Need for hospitalization in the intensive care unit | | | | 1  2 | | | |
| #176 | Were you returning from abroad when when you got sick? | Yes  No | | | | 1  2 | | | |
| #177 | In the past 12 months, have you experienced any of the following symptoms? *(several possible answers)* | Fever (≥38 °C)  Sore throat  Runny nose  Cough  Shortness of breath  Chills  Vomiting  Nausea  Diarrhea  Headache  Skin rash  Conjunctivitis  Muscle aches  Joint pain  Loss of appetite  Loss of smell  Loss of taste  Fatigue  Other symptoms  Asymptomatic  If Other (please specify: | | | | 1  2  3  4  5  6  7  8  9  10  11  12  13  14  15  16  17  18  19  20  …………………… Cov10_other | | | |
| #178 | Have you been in contact with a suspected or confirmed case of COVID-19? | Yes  No  Don’t know | | | | 1  2  77 | | | |
| #179 | Has a family member tested positive for Covid-19? | Yes  No  Don’t know | | | | 1  2  77 | | | |
| #180 | Has a work colleague tested positive for Covid-19? | Yes  No  Don’t know | | | | 1  2  77 | | | |
| #181 | Have you participated in a group gathering in which one or more cases were detected (festive meal, family meal, etc.)? | Yes  No  Don’t know | | | | 1  2  77 | | | |
| #182 | Did you have any prevention actions in place? (several possible answers) | Hand washing or hand disinfection  Don't shake hands  Cough into your elbow  Maintain a safe distance  Wearing a mask  None | | | | 1  2  3  4  5  6 | | | |
| What has been the impact of the health crisis on your daily life? | | | | | | | | | |
| #183 | Social (social support, loss of social ties, isolation) | Positive  Negative  Both  Neither of the two  Don’t know | | | | 1  2  3  4  77 | | | |
| #184 | Psychological (well-being, relaxation, joy, sadness, distress, stress, etc.) | Positive  Negative  Both  Neither of the two  Don’t know | | | | 1  2  3  4  77 | | | |
| #185 | Economic (employment, increase in income, retraining, versatility, loss of work, increased or reduced income) | Positive  Negative  Both  Neither of the two  Don’t know | | | | 1  2  3  4  77 | | | |
| #186 | Family (family reunification, family breakdown, etc.) | Positive  Negative  Both  Neither of the two  Don’t know | | | | 1  2  3  4  77 | | | |
| #187 | Health (absence of appearance of common diseases due to the prevention actions in place, diseases with clinical complications, etc.) | Positive  Negative  Both  Neither of the two  Don’t know | | | | 1  2  3  4  77 | | | |
| #188 | Are you vaccinated? | Yes  No  Don’t want to answer | | | | 1  2  88 | | | |
| #189 | Product name of the vaccine | Pfizer/BioNTech  Moderna  Janssen  AstraZeneca  Other | | | | 1  2  3  4  5 | | | |
|  |  | If Other (please specify): | | | | …………………… | | | |
| #190 | First injection date: | Date | | | | _ _ _ _ _ _ | | | |
| #191 | Second injection date | Date | | | | _ _ _ _ _ _ | | | |
| **Mental health** | | | | | | | | | |
|  | **Question** | **Answer** | | | | | | | |
| #192 | We are going to ask you sensitive questions related to mental health. Do you accept to answer it? | Yes  No | | | | 1  2 | | | |
| How often have you been bothered by the following over the past 2 weeks? | | | | | | | | | |
| #193 | - Little interest or pleasure in doing things? | Not at all  Several days  More than half the days  Nearly every day  Don’t know  Don’t want to answer | | | | 1  2  3  4  77  88 | | | |
| #194 | - Feeling down, depressed, or hopeless? |  |  |  |  |  |  |  |  |
| #195 | - Trouble falling or staying asleep, or sleeping too much? |  |  |  |  |  |  |  |  |
| #196 | - Feeling tired or having little energy? |  |  |  |  |  |  |  |  |
| #197 | - Poor appetite or overeating? |  |  |  |  |  |  |  |  |
| #198 | - Feeling bad about yourself — or that you are a failure or have let yourself or your family down? |  |  |  |  |  |  |  |  |
| #199 | - Trouble concentrating on things, such as reading the newspaper or watching television? |  |  |  |  |  |  |  |  |
| #200 | - Moving or speaking so slowly that other people could have noticed? Or so fidgety or restless that you have been moving a lot more than usual? |  |  |  |  |  |  |  |  |
| #201 | - Thoughts that you would be better off dead, or thoughts of hurting yourself in some way? |  |  |  |  |  |  |  |  |
| #202 | If you checked off any problems, how difficult have these problems made it for you to do your work, take care of things at home, or get  along with other people? | Not difficult at all  Somewhat difficult  Very difficult  Extremely difficult  Don’t know Don’t want to answer | | | | 1  2  3  4  77  88 | | | |
| **Sexual health** | | | | | | | | | |
|  | **Question** | **Answer** | | | | | | | |
| #203 | We are going to ask you sensitive questions related to sexual health. Do you accept to answer it? | Yes  No | | | | 1  2 | | | |
| #204 | Have you ever had sexual intercourse? | Yes  No  Don’t want to answer | | | | 1  2  88 | | | |
| #205 | How old were you when you first had sexual intercourse? | Age in Years  Don’t remember  Don’t want to answer | | | | _ _  77  88 | | | |
| #206 | On the first time that you had sexual intercourse, did you or your partner use a condom against sexually transmitted infections? | Yes  No  Don't remember  Don’t know  Don’t want to answer | | | | 1  2  3  77  88 | | | |
| #207 | When you have a casual sexual intercourse, do you use a condom against sexually transmitted infections? | Yes  No  Sometimes  Don’t have any casual sexual intercourse  Don’t want to answer | | | | 1  2  3  4  88 | | | |
| #208 | Have you ever had a disease/ infection which you got through sexual contact? | Yes  No  Don’t know  Don’t want to answer | | | | 1  2  77  88 | | | |
| **Female health** | | | | | | | | | |
|  | **Question** | **Answer** | | | | | | | |
| #209 | We are going to ask you sensitive questions related to women health. Do you accept to answer it? | Yes  No | | | | 1  2 | | | |
| #210 | At what age did you have your first period? | Years  Don’t know  Don’t want to answer | | | | _ _  77  88 | | | |
| #211 | Are you currently taking contraception? | Yes  No  Don’t know  Don’t want to answer | | | | 1  2  77  88 | | | |
| #212 | If Yes, which one? | Contraceptive pill  IUD  Other | | | | 1  2  3 | | | |
| #213 | Are you menopausal? | Yes  No  Don’t know  Don’t want to answer | | | | 1  2  77  88 | | | |
| #214 | If Yes, at what age did you have your menopause? | Years  Don’t know  Don’t want to answer | | | | _ _  77  88 | | | |
| #215 | How many times have you been pregnant? | Number of pregnancies  Don’t know  Don’t want to answer | | | | _ _  77  88 | | | |
| #216 | How many of these pregnancies were miscarriages? | Number of miscarriages  Don’t know  Don’t want to answer | | | | _ _  77  88 | | | |
| **Physical measurements** | | | | | | | | | |
| **Blood pressure** | | | | | | | | | |
|  | **Question** | **Answer** | | | | | | | |
| #217 | Interviewer ID |  | | | | _ _ | | | |
| #218 | Device ID for blood pressure |  | | | | _ _ | | | |
| #219 | Cuff size used | Small  Medium  Large  Extra-large | | | | 1  2  3  4 | | | |
| #220 | Reading 1 | Systolic ( mmHg) | | | | _ _ _ | | | |
|  |  | Diastolic (mmHg) | | | | _ _ _ | | | |
| #221 | Reading 2 | Systolic ( mmHg) | | | | _ _ _ | | | |
|  |  | Diastolic (mmHg) | | | | _ _ _ | | | |
| #222 | Reading 3 | Systolic ( mmHg) | | | | _ _ _ | | | |
|  |  | Diastolic (mmHg) | | | | _ _ _ | | | |
| #223 | During the past two weeks, have you been treated for raised blood pressure with drugs (medication) prescribed by a doctor or other health worker? | Yes  No | | | | 1  2 | | | |
| **Height and weight** | | | | | | | | | |
|  | **Question** | **Answer** | | | | | | | |
| #224 | For women: Are you pregnant? | Yes  No | | | | 1 *If Yes, go to #231*  2 | | | |
| #225 | Interviewer ID |  | | | | _ _ | | | |
| #226 | Device IDs for height and weight | Height | | | | _ _ | | | |
|  |  | Weight | | | | _ _ | | | |
| #227 | Height | in Centimetres (cm) | | | | _ _ _ . _ | | | |
| #228 | Weight  *If too large for scale 666.6* | in Kilograms (kg) | | | | _ _ _ . _ | | | |
| **Waist** | | | | | | | | | |
|  | **Question** | **Answer** | | | | | | | |
| #229 | Device ID for waist |  | | | | _ _ | | | |
| #230 | Waist circumference | in Centimetres (cm) | | | | _ _ _ . _ | | | |
| **Skin color measurement** | | | | | | | | | |
|  | **Question** | **Answer** | | | | | | | |
| #231 | Reading n°1 |  | | | |  | | | |
| #232 | Reading n°2 |  | | | |  | | | |
| #233 | Reading n°3 |  | | | |  | | | |
| #234 | Reading n°4 |  | | | |  | | | |
| **Blood glucose** | | | | | | | | | |
|  | **Question** | **Answer** | | | | | | | |
| #235 | During the past 12 hours have you had anything to eat or drink, other than water? | Yes  No | | | | 1  2 | | | |
| #236 | Technician ID |  | | | | _ _ | | | |
| #237 | Device ID |  | | | | _ _ | | | |
| #238 | Time of day blood specimen taken (24 hour clock) | Hours : minutes | | | | _ _ : _ _  hrs mins | | | |
| #239 | Fasting blood glucose | mg/dl | | | | _ _ _ _ | | | |
| **#**240 | Today, have you taken insulin or other drugs (medication) that have been prescribed by a doctor or other health worker for raised blood glucose? | Yes  No | | | | 1  2 | | | |
